# Contextualizing future maternal RSV vaccination acceptance and trust among pregnant and lactating women in Kenya: A latent class analysis

**DOI:** 10.1101/2025.03.24.25324585

**Authors:** Molly A. Sauer, Berhaun Fesshaye, Emily Miller, Jessica Schue, Prachi Singh, Rose Jalang’o, Joyce Nyiro, Christine Karanja-Chege, Rosemary Njogu, Fred Were, Ruth Karron, Rupali J. Limaye

**Affiliations:** Department of International Health, Johns Hopkins Bloomberg School of Public Health, Baltimore, Maryland, USA; National Vaccines and Immunization Program, Ministry of Health, Nairobi, Kenya; Kenya Medical Research Institute-Wellcome Trust Research Programme, Kilifi, Kenya; Department of Paediatrics and Child Health, Kenyatta University, Nairobi, Kenya; Jhpiego, Nairobi, Kenya; Department of Paediatrics & Child Health, University of Nairobi, Nairobi, Kenya; Department of Global and Community Health, George Mason University, Fairfax, Virginia, USA

**Keywords:** maternal vaccination, respiratory syncytial virus, vaccine acceptance, latent class analysis, Kenya

## Abstract

Maternal vaccination, or vaccination in pregnancy, offers a critical opportunity to provide protection to pregnant women and simultaneously confer passive immunity to infants in the first months of life, when infections are particularly serious and their immune systems are still developing. Respiratory syncytial virus (RSV) is one such serious infectious disease for newborns, but a newly approved and recommended vaccine for respiratory syncytial virus has been designed to be given to pregnant women to protect their newborns from severe RSV disease when they are most vulnerable. While maternal vaccination has been used for tetanus, pertussis, influenza, COVID-19, and other diseases, vaccination in pregnancy can present unique challenges related to hesitancy and delivery, particularly in lower-resourced settings. Using data from a cross-sectional survey of 400 pregnant and lactating women in Nakuru and Mombasa Counties in Kenya, we examined knowledge, attitudes, and beliefs related to maternal vaccination and RSV through a latent class analysis. We identified two distinct archetypes among study subjects, maternal vaccine questioners and maternal vaccine acceptors, with notable differences in perceived vaccination-enabling social norms and use of private health facilities between the two groups. This respondent-driven approach to identify groups that may require different communication strategies can help to shape efforts to target preparations for future RSV vaccine introduction in Kenya and inform tailored health promotion strategies to support informed, confident maternal vaccination decision-making among providers, communities, and pregnant women.

## Background

Lower respiratory tract infections (LRTI) are second only to prematurity in contributing to global childhood mortality, responsible for an estimated 725,000 deaths among children under 5 in 2021—about one in every seven under-5 deaths (1). Among the leading causes of LRTI hospitalizations and deaths are viral causes, like respiratory syncytial virus (RSV) and influenza, and bacteria, such as *Streptococcus pneumoniae* (pneumococcus) and *Haemophilus influenzae* type b (Hib) (2). Improvements in case management, nutrition, access to antibiotics and oxygen, and water, sanitation, and hygiene have helped reduce the burden of LRTI, and vaccines for some of the most common and serious causes of childhood LRTI have also played a significant role. Today, pneumococcal, Hib, influenza, and pertussis vaccines help protect infants and young children from serious illness; in just the last few years, a maternal vaccine and immunoprophylaxis for RSV have been added to the global immunization portfolio as interventions to reduce this leading cause of LRTI hospitalizations and deaths (2–4).

### RSV disease and global burden

RSV is a leading cause of respiratory illness and pneumonia morbidity and mortality (4,5). While RSV infection in healthy adults generally presents as a mild respiratory illness (6), it is particularly pervasive and dangerous for young infants. It is estimated that every child has been infected by RSV by 2 years of age (7), with incidence of severe RSV disease peaking in the second month of life (8). Seasonality varies, with some areas experiencing year-round transmission and others with defined seasonal peaks; still others have indeterminate or unknown seasonal patterns in the absence of etiologic data for LRTI (9,10).

A systematic analysis by Li, et al (11) estimated that RSV was associated with more than 30 million LRTI episodes, 3.6 million hospital admissions, and more than 100,000 deaths among children under 5 in 2019. Li and colleagues estimated 6.6 million RSV-associated LRTI episodes among infants 0-6 months of age (1 in every 5 LRTI episodes in children under 5); almost 40% of RSV hospitalizations (1.4 million) and almost half of RSV deaths occurred in this age group. Infants who are born preterm (<37 weeks gestation) are at particularly high risk of serious RSV disease, but on a population basis, most infants aged 0-6 months with severe RSV LRTI are healthy full-term infants without identifiable risk factors (5).

While RSV affects children worldwide, it is far from democratic; an estimated 95% of RSV- associated LRTI episodes and 97% of deaths occur in low- and middle-income countries (3,11–13). In Kenya, RSV infection is associated with a substantial burden of disease for young children, with as many as 365,000 episodes among children under 5 years of age each year (11,14). Recent analyses estimated RSV was responsible for more than a third (36.1% [95% CI: 32.1, 41.0]) of severe pneumonia cases among children under 5 in Kenya, a greater contributor by far than other viral, bacterial, or fungal pathogens (2).

### RSV prevention through passive immunization

RSV disease is most serious in the first months of life when infant immune systems are unable to generate antibodies while also mounting an immune response to acute infection (3,7,12,15). Thus, passive immunization strategies have been a priority to provide protection to infants.

Unlike most vaccines, which use non-replicating or weakened replicating versions of pathogens to train infants’ immune systems to recognize and build targeted antibodies without first being infected with the disease, passive immunization confers immunity by providing pathogen-specific antibodies to the infant for immediate—albeit relatively short-lived—protection from the target pathogen (16–18). Leveraging passive immunization to protect the newborn is not a new concept and has been used for a range of other diseases such as tetanus, for example. Passive immunization is an essential strategy to intervene at the most vulnerable period when RSV disease is most severe. Fortunately, significant strides have recently been made in this area with two new interventional tools approved in the last three years to prevent severe RSV disease in infants through vaccination in pregnancy and immunoprophylaxis with monoclonal antibodies in infancy (3,12).

A single-dose extended half-life RSV monoclonal antibody (mAb), nirsevimab, was approved in 2022 and confers about 5-6 months protection when administered in infants (3,19). Nirsevimab has been rolled out in several high-income settings but remains prohibitively expensive for most low- and middle-income country (LMIC) health systems, and substantial supply shortages have already constrained its use even in high-income settings (3,20,21). For now, mAbs are poised to remain a tool for wealthier countries and out of reach of children in LMICs (15).

Issues of equitable access to mAbs give added weight to the importance and opportunity of maternal vaccination. By administering an RSV vaccine to the mother during pregnancy, antibodies are passed transplacentally to the fetus in utero and to the infant via breastmilk. While maternal antibodies wane over time, this mechanism is important and effective in protecting children during the first six months of life when RSV disease is most serious (3,22). The first maternal RSV vaccine, RSVpreF, was developed by Pfizer and approved by regulatory authorities in the USA in 2023 and is administered as a single dose to pregnant women in the third trimester (12,23). In clinical trials, maternal RSVpreF demonstrated a vaccine efficacy of 70% in preventing severe RSV-LRTI in the first 6 months and over 80% in the first 90 days (3,22).

### Enabling maternal vaccine acceptance and uptake

The availability of a safe, effective, and approved maternal vaccine for RSV is an essential and exciting breakthrough, and the path to widespread uptake in lower-resourced settings with the highest burden of disease is winding. Some key challenges include vaccine affordability and availability, delivery platforms for pregnant women, RSV seasonality, vaccine hesitancy, and the continued need for post-licensure surveillance and studies (3,12).

In September 2024, the World Health Organization (WHO) Strategic Advisory Group of Experts on Immunization (SAGE) recommended that all countries introduce maternal RSV vaccines and/or nirsevimab to prevent severe RSV disease in infants (3). RSVpreF has been approved by regulatory authorities in several high- and upper-middle-income countries and has been introduced in the USA, Argentina, and a handful of others (12). Additionally, Gavi, the Vaccine Alliance—which provides co-financing and other support to eligible LMICs to introduce and sustain vaccines in the national immunization program—included RSV vaccines in their 2024 Vaccine Investment Strategy for the 2026-2030 period, indicating intent to support maternal RSV vaccine introduction once a suitable candidate is approved and WHO-prequalified (24).

Current maternal vaccines like tetanus toxoid-containing vaccines (TTCV) and COVID-19 vaccines are administered during antenatal care (ANC) visits. However, uptake of ANC services is variable; in Kenya, 66% of pregnant women had at least four ANC visits during their pregnancy and fewer still had the recommended eight ANC visits. About one in 10 pregnant women (9.3%) did not have their first ANC visit until the third trimester, the time at which RSVpreF would need to be given. In two counties, Garissa and Mandera, one in five women did not have any ANC prior to delivery (25).

The varied seasonality of RSV in different regions of the world further complicates maternal RSVpreF vaccination guidelines and recommendations. Where transmission is year-round or indeterminate—including most tropical and subtropical regions—maternal vaccination policies and practices are likely to be clearer, with year-round vaccination approaches recommended (3). Whether a woman is eligible to be vaccinated will be less contingent on what time of year she is due to deliver. In settings with well-defined and documented seasonal RSV patterns, maternal vaccination strategies may focus on offering the vaccine to pregnant women who are approaching delivery only in the lead-up to or during RSV peak seasons; however, year-round RSV vaccination may still be more feasible than seasonal strategies in these settings (3,12,26). Settings with seasonal patterns and co-availability of maternal vaccines and mAbs—largely limited to high income settings, given the high cost and low availability of nirsevimab—may consider a combined approach to close immunity gaps in infants born outside of the RSV season to mothers ineligible for the vaccine.

Broadly, maternal vaccine acceptance is an important consideration and vaccine hesitancy— delaying or refusing a recommended vaccine—hinders the impact of these interventions. TTCVs have been given in pregnancy for decades in LMICs, but shifting COVID-19 vaccine recommendations for pregnant women led to confusion and lower uptake in many settings, including Kenya (27). Maternal RSV vaccines will be introduced in the shadow of COVID vaccine rollout, and it remains to be seen how communities, providers, and pregnant women will perceive these new vaccines. Systems and communities familiar with administering TTCV and COVID-19 vaccines at any time in pregnancy will also need to navigate the narrow window for RSV vaccination, as the maternal RSV vaccine is recommended only in the third trimester.

### Study purpose and approach

Latent class analysis (LCA) is a subject-centered statistical approach to identify underlying, unobserved (“latent”) groups (“classes”) that exist among subjects based on their responses to a set of measured or observed parameters. LCA allows for analyses to be shaped by respondents rather than ad hoc or a priori groupings established by researchers or others. LCA can examine patterns in individuals’ responses to determine if there are organically forming groupings amongst subjects based on those patterns. In this study, we examined how respondents understand, perceive, and feel about maternal vaccination and RSV disease in order to identify and describe emergent archetypes. We then assigned probabilities of each response based on membership in a given class, and on individual subjects’ probability of being assigned to a given class based on their response pattern.

Kenyan scientific and policy officials are already exploring the potential for introducing maternal RSV vaccines. Through LCA, and a brief exploration of sociodemographic factors associated with class membership, we aimed to identify discrete archetypes or classes that might exist among pregnant (and recently pregnant) women. We further sought to describe shared characteristics or perspectives within classes as well as the characteristics and perspectives that may distinguish each class.

## Methods

To explore underlying archetypes among pregnant and lactating women in Nakuru and Mombasa Counties, in Kenya, and what factors may be associated with each class, we conducted an LCA and simple logistic regression using data collected through a cross-sectional survey under the Maternal Immunization Readiness Initiative (MIRI) in July-September 2022 (28). This paper uses the term “pregnant women” in consultation with local partners on culturally appropriate language and to reflect respondents self-identified gender; all respondents self-identified as “women” when asked for their gender (woman, man, other). Although most people who are or can get pregnant are cisgender women who were born and identify as female, these topics are also relevant to the experiences of transgender men and other gender diverse people who may have the capacity to become pregnant.

### Study population

Survey data were collected at 20 health facilities across Nakuru and Mombasa Counties, Kenya, as part of MIRI’s broad multi-method study on demand for maternal COVID-19, RSV, and group B streptococcus vaccines among pregnant and lactating women, community members, health providers, and policymakers (27,29–34). These two counties and the facilities selected were selected in consultation with study partners and Ministry of Health (MOH) officials to maximize geographic and sociodemographic diversity as well as study feasibility. Nakuru County is a relatively rural county located west of Nairobi. While home to one of Kenya’s largest cities (Nakuru), the county is predominantly rural, and most residents are involved in agriculture-related industries. Conversely, Mombasa County is a geographically small but densely populated urban county located on Kenya’s Indian Ocean coast near the border with Tanzania to the south. The county comprises Mombasa city. Facilities ranged from level 2 facilities (community health outposts) to level 5 facilities (large referral hospitals).

### Source data: Cross-sectional survey of pregnant and lactating women

For the cross-sectional survey of pregnant and lactating women, we developed a survey instrument drawing from the Parent Attitudes about Childhood Vaccines (PACV) questionnaire, a validated and widely used survey tool developed by Opel, et al. (35–37) to measure parental hesitancy related to pediatric vaccination, and the Vaccine Hesitancy Scale (VHS) developed by the WHO SAGE Working Group on Vaccine Hesitancy to inform immunization program and policy interventions (38). We adapted these scales to consider vaccination in pregnancy, asking about perspectives on both mother and infant, for example. Instruments were developed in English; we translated the survey instrument into Swahili and piloted the translated version to confirm translation accuracy and appropriateness. Data collectors underwent a multi-day training on relevant scientific and socio-behavioral concepts, ethical research practice, and data management.

We enrolled pregnant or lactating women across 20 facilities using a convenience sampling approach; target enrollment varied by facility based on facility catchment area size and typical facility attendance. Data collection began on 25 July 2022 and ended on 30 September 2022. Potential participants were approached consecutively within health facilities, typically when arriving for antenatal, post-natal, or newborn clinics, or upon referral from health facility personnel. They were screened for eligibility and provided information about the study.

Individuals meeting the inclusion criteria (at least 18 years of age, able to provide consent, and either currently in the second or third trimester of pregnancy or currently breastfeeding) and interested in participating in the study were administered oral consent before beginning the survey. Surveys were conducted in English or Swahili, by trained data collectors using tablets; this included a 10-second video showing a child with classic RSV symptoms at the start of the survey. Data and consent were collected and stored on encrypted servers using REDCap (39,40).

The parent study received ethical approval from institutional review boards (IRB) at the Johns Hopkins Bloomberg School of Public Health (JHSPH), the Kenya Medical Research Institute (KEMRI), and the National Commission for Science, Technology & Innovation (NACOSTI). Data collection was conducted with the approval of the Kenya Ministry of Health and county health authorities in Nakuru and Mombasa counties.

Survey results were initially summarized with descriptive statistics and a simple logistic regression examining respondent characteristics, relevant social and behavioral constructs, and associations with vaccine hesitancy. These findings are presented in a previously published article by Limaye, et al (29). The study identified primigravida status—first pregnancy/child—as the only factor statistically significantly associated with higher vaccine hesitancy among pregnant or lactating women, in a simple logistic regression (29).

### Latent class analysis

#### Variables considered in the model

For the LCA, we considered 12 survey items related to knowledge, attitudes, and beliefs (KAB) about RSV disease, RSV vaccines in pregnancy, and maternal vaccines generally. Responses were measured on a four-point Likert scale; knowledge-related questions were measured on a five-point scale, including a “don’t know” option. Variables assessing respondents’ perceptions of disease risk and seriousness included measures of perceived prevalence of RSV, perceived susceptibility, and perceived severity of RSV disease for the mother and baby. Two items assessed perceived safety of vaccines in pregnancy for the mother and baby. Two items assessed perceived benefits or effectiveness of maternal vaccines for the mother and baby.

Two items measured social norms related to vaccination in pregnancy. One item assessed perceived self-efficacy to get a vaccine in pregnancy and one item examined perceived barriers to care or vaccination. Survey items and response options are presented in Supplementary Table 1 (S1 Table).

Responses were dichotomized into “strongly agree or agree” and “disagree or strongly disagree”. For the four knowledge-related questions (perceived prevalence, perceived susceptibility, and perceived severity for mother and baby), we dichotomized responses to “strongly agree or agree” and “disagree, strongly disagree, or don’t know.”

#### Model testing and selection

Using Mplus version 8.9 (Los Angeles, CA, USA) (41), we compared models with two to five classes to determine best fit and select a final model. We explored measures of relative model fit: the Akaike Information Criteria (AIC) and Bayesian Information Criteria (BIC), with lower values indicating better model fit, and the Lo-Mendell-Rubin likelihood ratio test (LMR-LRT) and bootstrapped likelihood ratio test (BLRT), with statistically significant values indicating an improvement in model fit comparing the *k* class model to the *k-1* class model. We also explored additional model characteristics, including measures of absolute model fit (Pearson chi-square and likelihood ratio chi-square), entropy, and assignment probabilities to identify and navigate any issues with our model. Prior to conducting the analysis, we determined that we would prioritize BIC, LMR-LRT, and BLRT for final model selection. We conducted subsequent analyses in Stata version 18 (College Station, TX, USA) (42).

### Logistic regression

Following the LCA and identification of class conditional probabilities, we explored sociodemographic factors associated with class membership through simple and adjusted logistic regression. Our dependent variable was class membership and independent variables included key sociodemographic characteristics collected in the cross-sectional survey (i.e., age, county, education level, primigravida status, facility type). Results are presented as odds ratios and p-values (p ≤ 0.05 indicates statistical significance). Data cleaning and analysis was conducted using Stata version 18 (College Station, TX, USA) (42).

## Results

### Sample description

Survey responses were evenly split between the two counties. Of the 400 individuals enrolled and surveyed, 269 (67.2%) were 18-29 years of age and 131 (32.8%) were 30-44 years of age. This was the first pregnancy or child for 65% (260) of respondents; three-quarters (299, 74.8%) of respondents were breastfeeding. About 17% (67) of respondents had previously delayed or refused a vaccine during pregnancy, and about 24% (97) had previously delayed or refused a vaccine when they were not pregnant. About one in ten (10.5%) indicated a preference for illness-acquired immunity compared to vaccine-induced immunity. Side effects were the highest ranked concern among respondents, with more than half (51.2%) rating side effects as their top concern (1 out of 7) compared to vaccine ingredients, costs, availability, what others say about the vaccine, family input, and health worker recommendations.

### Respondent archetypes

We examined 2-, 3-, 4-, and 5-class models based on 12 items measuring respondent KAB related to maternal vaccination and RSV. We noted the lowest BIC (3300.74) in the 3-class model; however, the LMR-LRT was not significant (p = 0.221), indicating that the 3-class model did not significantly improve upon the 2-class model. This pairing of a low BIC and non-significant LMR-LRT may also indicate that the 3-class model may be overfitting the data. The 2-class model had only a marginally higher BIC (3302.21) and a statistically significant LMR-LRT (p<0.001). Based on fit statistics and parsimony, we determined the 2-class model was optimal (Table 1).

**Table 1.** Goodness-of-fit statistics for LCA models.

| Model | Log-likelihood | AIC | BIC | Entropy | LMR-LRT($p$) | BLRT($p$) |
| --- | --- | --- | --- | --- | --- | --- |
| 2 class | -1576.21 | 3202.42 | 3302.21 | 0.72 | 0.000 | 0.000 |
| 3 class | -1536.53 | 3149.06 | 3300.74 | 0.95 | 0.221 | 0.000 |
| 4 class | -1519.70 | 3141.41 | 3344.97 | 0.97 | 0.163 | 0.020 |
| 5 class | -1504.51 | 3137.03 | 3392.48 | 0.83 | 0.328 | 0.000 |

*AIC = Akaike information criterion; BIC = Bayesian information criterion; LMR-LRT = Lo-Mendell-Rubin likelihood ratio test; BLRT = Bootstrapped likelihood ratio test*

Of the 400 pregnant or lactating women surveyed, 95 (23.8%) were classified as “questioners” (class 1) and 305 (76.2%) were classified as “acceptors” (class 2) based on responses to the 12 KAB variables included in the model (Fig 1).

**Fig 1.**
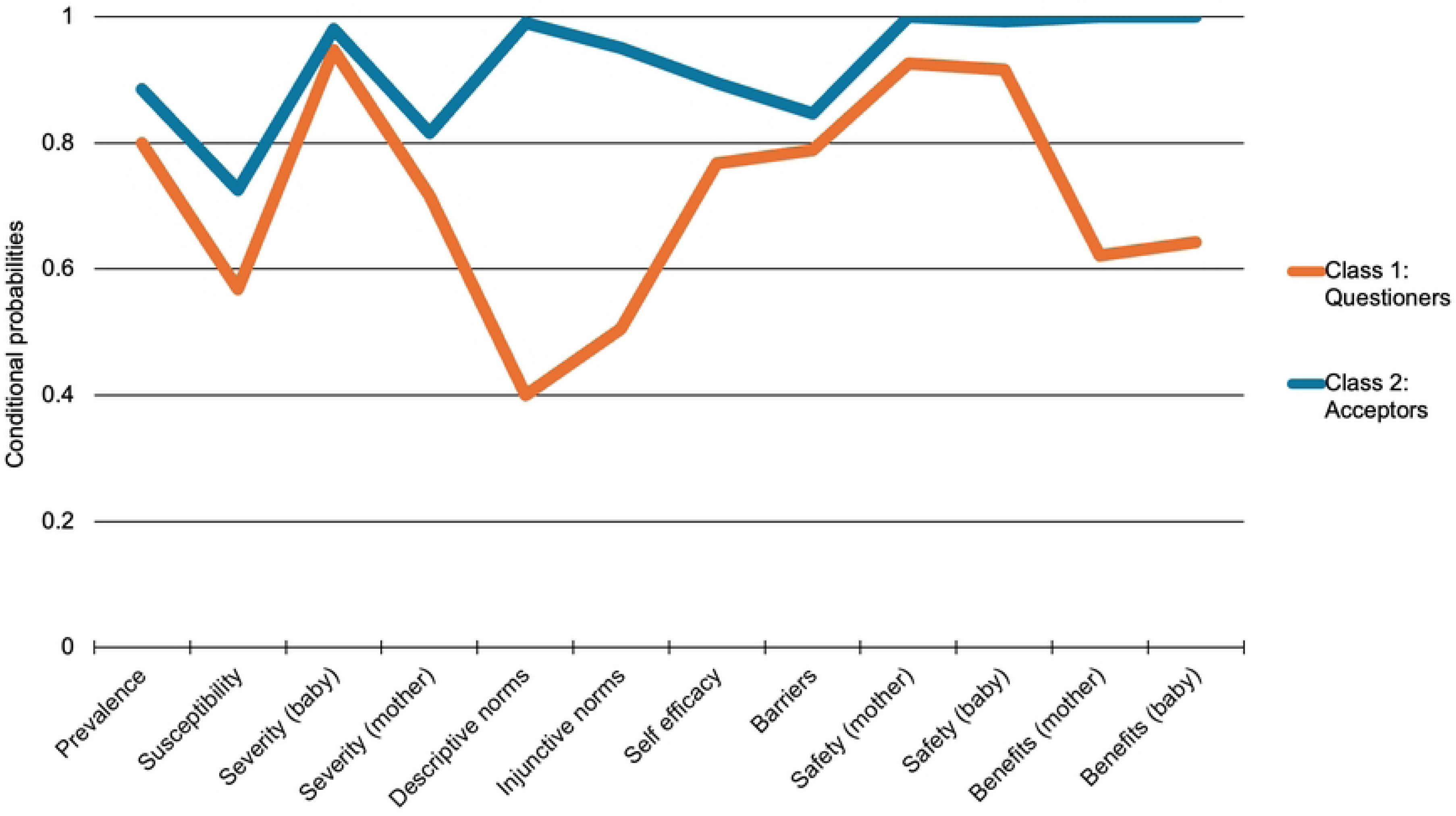
Conditional probabilities^a^ for each variable, by class membership. ^a^Probability of affirmative (agree/strongly agree) response to each variable, given class membership; variables described in Table 2.

**Table 2.** Maternal vaccination-related KAB probabilities (displayed as percentages) by latent class membership.

| Variable | Class 1:<br>Questioners<br>(n=95, 23.8%) | Class 2:<br>Acceptors<br>(n=305, 76.2%) | p |
| --- | --- | --- | --- |
| The majority of babies <2 years old get RSV. *[perceived prevalence]* |  |  | 0.034* |
| Strongly agree or agree | 76 (80.0%) | 270 (88.5%) |  |
| Disagree, strongly disagree, or don't know | 19 (20.0%) | 35 (11.5%) |  |
| I worry that my baby could get RSV. *[perceived susceptibility]* |  |  | 0.004* |
| Strongly agree or agree | 54 (56.8%) | 221 (72.5%) |  |
| Disagree, strongly disagree, or don't know | 41 (43.2%) | 84 (27.5%) |  |
| I believe RSV is dangerous for babies. *[perceived severity - infant]* |  |  | 0.086 |
| Strongly agree or agree | 90 (94.7%) | 299 (98.0%) |  |
| Disagree, strongly disagree, or don't know | 5 (5.3%) | 6 (2.0%) |  |
| I believe RSV is dangerous for pregnant women or women who have recently given birth. *[perceived severity - mother]* |  |  | 0.035* |
| Strongly agree or agree | 68 (71.6%) | 249 (81.6%) |  |
| Disagree, strongly disagree, or don't know | 27 (28.4%) | 56 (18.4%) |  |
| If there was an MOH approved maternal vaccine for RSV, the majority of my pregnant friends and family would get it. *[descriptive social norms]* |  |  | <0.001** |
| Strongly agree or agree | 38 (40.0%) | 302 (99.0%) |  |
| Disagree or strongly disagree | 57 (60.0%) | 3 (1.0%) |  |
| If there was an approved maternal vaccine for RSV, the majority of my friends and family would encourage me to get it. *[injunctive social norms]* |  |  | <0.001** |
| Strongly agree or agree | 48 (50.5%) | 290 (95.1%) |  |
| Disagree or strongly disagree | 47 (49.5%) | 15 (4.9%) |  |
| I have some control over whether or not I get vaccines during my pregnancy. *[self-efficacy]* |  |  | 0.002* |
| Strongly agree or agree | 73 (76.8%) | 273 (89.5%) |  |
| Disagree or strongly disagree | 22 (23.2%) | 32 (10.5%) |  |
| If I need to visit a health facility for an appointment or a vaccine, I can easily go to that health facility. *[barriers]* |  |  | 0.198 |
| Strongly agree or agree | 75 (78.9%) | 258 (84.6%) |  |
| Disagree or strongly disagree | 20 (21.1%) | 47 (15.4%) |  |
| I am confident that vaccines recommended for me during pregnancy are safe for me. *[safety - mother]* |  |  | <0.001** |
| Strongly agree or agree | 88 (92.6%) | 305 (100.0%) |  |
| Disagree or strongly disagree | 7 (7.4%) | 0 (0.0%) |  |
| I am confident that vaccines recommended for me during pregnancy are safe for my baby. *[safety - infant]* |  |  | <0.001** |
| Strongly agree or agree | 87 (91.6%) | 303 (99.3%) |  |
| Disagree or strongly disagree | 8 (8.4%) | 2 (0.7%) |  |
| If a new vaccine were approved for use among pregnant women, I trust that the vaccine would protect me. [*benefits - mother*] |  |  | <0.001** |
| Strongly agree or agree | 59 (62.1%) | 305 (100.0%) |  |
| Disagree or strongly disagree | 36 (37.9%) | 0 (0.0%) |  |
| If a new vaccine were approved for use among pregnant women, I trust that the vaccine would protect the fetus/baby. [*benefits - baby*] |  |  | <0.001** |
| Strongly agree or agree | 61 (64.2%) | 305 (100.0%) |  |
| Disagree or strongly disagree | 34 (35.8%) | 0 (0.0%) |  |
| * $p \leq 0.05$; ** $p \leq 0.001$ | | | |

Table 2 presents conditional probabilities for each KAB item included in the model, given class membership—the likelihood of observing a specific response within that class, assuming all variables are independent. Differences between the two classes were statistically significant for 10 variables: perceived RSV prevalence, perceived RSV susceptibility, perceived RSV severity for pregnant women, injunctive and descriptive social norms, self-efficacy, and vaccine safety of the vaccine for mother, perceived safety of the vaccine for the baby, perceived benefits/effectiveness for the mother, and perceived benefits/effectiveness for the baby. Using the 2-class model, perceived barriers to vaccination in pregnancy and perceived RSV severity for the baby were not statistically significantly different between the two classes.

#### Sociodemographic characteristics

Respondents were asked to provide information about their age, number of children under 18, whether they were pregnant or lactating, and their education level. We also documented whether they were enrolled while attending a public or private facility. We generated a parity variable based on responses to pregnancy/lactating status and number of children under 18; we classified respondents as primigravida (first pregnancy or child) or multigravida (not first pregnancy or child). Examining these sociodemographic factors, we noted a statistically significant difference in health facility type, given class membership (Table 3). Private facility use was more common among class 1 (“questioners”) than class 2 (“acceptors”), serving as a possible proxy for socioeconomic status. About 15% of “questioners” attended private health facilities and 85% were surveyed while attending a public (MOH) health facility, compared to just under 7% of “acceptors” seeking care at private facilities and 93% at public clinics (p = 0.013).

**Table 3.** Sociodemographic characteristics, information sources, influences, and other KAB factors by latent class membership.

| Variable | Total<br>(n=400) | Class 1:<br>Questioners<br>(n=95, 23.8%) | Class 2:<br>Acceptors<br>(n=305, 76.2%) | p |
| --- | --- | --- | --- | --- |
| **Sociodemographic characteristics** |  |  |  |  |
| Age |  |  |  | 0.436 |
| 18-29 | 269 (67.2%) | 67 (70.5%) | 202 (66.2%) |  |
| 30-44 | 131 (32.8%) | 28 (29.5%) | 103 (33.8%) |  |
| County |  |  |  | 0.411 |
| Nakuru | 200 (50.0%) | 44 (46.3%) | 156 (51.1%) |  |
| Mombasa | 200 (50.0%) | 51 (53.7%) | 149 (48.9%) |  |
| Highest education level |  |  |  | 0.709 |
| Less than primary school | 45 (11.2%) | 13 (13.7%) | 32 (10.5%) |  |
| Primary school | 130 (32.5%) | 31 (32.6%) | 99 (32.5%) |  |
| Secondary/high school | 121 (30.2%) | 25 (26.3%) | 96 (31.5%) |  |
| College/university or post-graduate | 104 (26.0%) | 26 (27.4%) | 78 (25.6%) |  |
| Pregnancy status |  |  |  | 0.990 |
| Breastfeeding | 299 (74.8%) | 71 (74.7%) | 228 (74.8%) |  |
| Pregnant (second trimester) | 45 (11.2%) | 11 (11.6%) | 34 (11.1%) |  |
| Pregnant (third trimester) | 56 (14.0%) | 13 (13.7%) | 43 (14.1%) |  |
| First pregnancy/child |  |  |  | 0.498 |
| No (multigravida) | 260 (65.0%) | 59 (62.1%) | 201 (65.9%) |  |
| Yes (primigravida) | 140 (35.0%) | 36 (37.9%) | 104 (34.1%) |  |
| Facility type |  |  |  | 0.013* |
| Public | 366 (91.5%) | 81 (85.3%) | 285 (93.4%) |  |
| Private | 34 (8.5%) | 14 (14.7%) | 20 (6.6%) |  |
| **Trusted information sources** |  |  |  |  |
| I trust the information that I have received from my health care provider about vaccines during pregnancy. |  |  |  | 0.933 |
| Strongly agree or agree | 392 (98.0%) | 93 (97.9%) | 299 (98.0%) |  |
| Disagree or strongly disagree | 8 (2.0%) | 2 (2.1%) | 6 (2.0%) |  |
| I trust the information that I have received from social media platforms (Facebook, WhatsApp) about vaccines during pregnancy. |  |  |  | 0.229 |
| Strongly agree or agree | 85 (21.2%) | 16 (16.8%) | 69 (22.6%) |  |
| Disagree or strongly disagree | 315 (78.8%) | 79 (83.2%) | 236 (77.4%) |  |
| I trust the information provided by scientists and doctors at universities and academic institutions about vaccines during pregnancy. |  |  |  | <0.001** |
| Strongly agree or agree | 339 (84.8%) | 70 (73.7%) | 269 (88.2%) |  |
| Disagree or strongly disagree | 61 (15.2%) | 25 (26.3%) | 36 (11.8%) |  |
| I trust the information that I have heard from the media (TV, radio, etc.) about vaccines during pregnancy. |  |  |  | 0.034* |
| Strongly agree or agree | 283 (70.8%) | 59 (62.1%) | 224 (73.4%) |  |
| Disagree or strongly disagree | 117 (29.2%) | 36 (37.9%) | 81 (26.6%) |  |
| **Vaccine hesitancy and acceptance measures** |  |  |  |  |
| Vaccines improve immunity. I believe it is better for me to develop my own immunity by getting sick rather than by getting a vaccine. |  |  |  | 0.694 |
| Strongly agree or agree | 42 (10.5%) | 11 (11.6%) | 31 (10.2%) |  |
| Disagree or strongly disagree | 358 (89.5%) | 84 (88.4%) | 274 (89.8%) |  |
| Have you ever delayed or decided not to get a recommended vaccine during pregnancy for reasons other than illness or allergy? |  |  |  | 0.026* |
| No | 333 (83.2%) | 72 (75.8%) | 261 (85.6%) |  |
| Yes or don't know | 67 (16.8%) | 23 (24.2%) | 44 (14.4%) |  |
| Thinking about your vaccine behavior when you were not pregnant, have you ever delayed or decided not to get a recommended vaccine for reasons other than illness or allergy? |  |  |  | 0.102 |
| No | 303 (75.8%) | 66 (69.5%) | 237 (77.7%) |  |
| Yes or don't know | 97 (24.2%) | 29 (30.5%) | 68 (22.3%) |  |
| If a new vaccine was approved for pregnant women, and your health care provider did recommend it, how likely would you get the new vaccine? |  |  |  | 0.030* |
| Very likely or likely | 370 (92.5%) | 83 (87.4%) | 287 (94.1%) |  |
| Unlikely or very unlikely | 30 (7.5%) | 12 (12.6%) | 18 (5.9%) |  |

\* $p \leq 0.05$ ; \*\* $p \leq 0.001$

No other sociodemographic factors differed significantly by class membership.

#### Trusted information sources and influences

Respondents were asked if they trust information on vaccination in pregnancy from each of four sources (their health care provider, social media, media (TV, radio, etc.), and university/academic doctors and researchers), with Likert-like response options from strongly agree to strongly disagree dichotomized to “strongly agree or agree” and “disagree or strongly disagree.” Trust in information received from health care providers was consistent and high across both classes (Table 3). Conversely, trust in information from academic scientists was significantly higher among “acceptors” than “questioners”; we also observed higher probability of trusting information from media sources among “acceptors” than “questioners.” Trust in information from social media was low in both classes.

### Factors associated with class membership

We conducted a simple logistic regression to examine variables that may be associated with class membership, with a focus on screening variables that could be observed or easily obtained by providers or immunization programs to guide vaccination efforts. Age, education level, county (as a proxy for rural or urban geographic area), primigravida/multigravida status, and health facility type were included as independent variables. The unadjusted and adjusted model, inclusive of all five factors, is shown in Table 4. Only health facility type (public vs. private) was statistically significantly associated with class membership—private facility users were nearly three times more likely to be “questioners” compared to public facility users, when adjusting for age, education, county, and primigravida status (OR: 2.986, p = 0.009).

**Table 4.** Logistic regression: Sociodemographic factors associated with membership in class 1 (“questioners”)

| Factor | Unadjusted model |  |  | Adjusted model |  |  |
| --- | --- | --- | --- | --- | --- | --- |
|  | OR | 95% CI | p | aOR | 95% CI | p |
| Age |  |  |  |  |  |  |
| 18-29 | Ref |  |  | Ref |  |  |
| 30-44 | 0.82 | (0.59, 1.35) | 0.436 | 0.814 | (0.47, 1.42) | 0.470 |
| County |  |  |  |  |  |  |
| Nakuru | Ref |  |  | Ref |  |  |
| Mombasa | 1.21 | (0.76, 1.93) | 0.411 | 1.17 | (0.73, 1.87) | 0.52 |
| Highest education level |  |  |  |  |  |  |
| Less than primary school | Ref |  |  | Ref |  |  |
| Primary school | 0.77 | (0.36, 1.65) | 0.502 | 0.75 | (0.34, 1.64) | 0.47 |
| Secondary/high school | 0.64 | (0.29, 1.40) | 0.264 | 0.58 | (0.25, 1.32) | 0.19 |
| College/university or post-graduate | 0.82 | (0.38, 1.79) | 0.620 | 0.54 | (0.22, 1.32) | 0.18 |
| First pregnancy/child |  |  |  |  |  |  |
| No (multigravida) | Ref |  |  | Ref |  |  |
| Yes (primigravida) | 1.18 | (0.73, 1.90) | 0.498 | 1.24 | (0.70, 2.14) | 0.460 |
| Facility type |  |  |  |  |  |  |
| Public | Ref |  |  | Ref |  |  |
| Private | 2.46 | (1.19, 5.09) | 0.015* | 2.99 | (1.32, 6.77) | 0.009 |

OR = odds ratio; aOR = adjusted odds ratio; CI = confidence interval; p = p-value (chi-square)

## Discussion

We identified two classes of pregnant or breastfeeding women based on 12 maternal vaccination KAB parameters. “Acceptors” (n=305, 76.2%) are more likely to express awareness and positive attitudes about RSV disease and maternal vaccination; “questioners” (n=95, 23.8%) are more likely to express negative or skeptical attitudes on RSV disease and maternal vaccination. While few studies were identified that used latent class analysis to explore underlying archetypes of maternal immunization beneficiaries, our findings generally align with the classification of pregnant women as confident or skeptical, although many identify a third class in the middle (i.e., confident, accepting, reluctant) (43,44). Interestingly, other studies have often indicated safety as a key driver of class distinction, whereas women in our study were largely in agreement about the safety of maternal vaccines for mother and baby regardless of class membership.

Risk perception is a key driver of vaccine behaviors, mediating the relationship between knowledge, awareness, and intentions. Individuals who perceive they are at high risk of infection and, if infected, are likely to have severe disease are more likely to accept a vaccine (45–49).

For pregnant women, perceived severity and susceptibility include considerations for the mother but also extend to—and often emphasize—the developing fetus and newborn (50,51). Lower perceived RSV prevalence and susceptibility among “questioners” is an important gap and one that can be addressed in advance of a vaccine introduction decision. RSV tends to have relatively non-specific symptoms similar to many LRTI-causing diseases of varying severity, and qualitative work on RSV in Kenya indicated that patients, community members, and even providers were unfamiliar with RSV as a unique and particularly severe disease rather than non-specific respiratory illness (29–31). There is an opportunity to begin training health providers to effectively discuss RSV with their patients, and to cascade this information to community health workers, before RSV vaccines are available to strengthen demand (among “acceptors”) and move the needle toward acceptance (for “questioners”).

In exploring sociodemographic factors associated with class membership, only facility type was statistically significantly associated with “questioner” status. Respondents who were enrolled at a private facility were far more likely to be “questioners” even when adjusting for other sociodemographic characteristics and KAB. While only 8% of study participants were enrolled in private facilities, our analysis indicated private sector health care seeking as a significant predictor of “questioner” class membership. Recent analyses indicate that about 45% of health facilities in Kenya are private (52)—however, our findings may indicate a meaningful difference between pregnant women accessing care through public facilities versus private facilities, including if and where the same providers may serve multiple facility types. We posit that there may be differences in socioeconomic status (SES) between private and public facility users that are not effectively captured in this analysis; future analyses could explore if facility type is an appropriate proxy for measures of SES and disentangle the effects of SES indicators and facility use as relating to maternal vaccination hesitancy.

Both groups had generally high trust in maternal vaccine safety for both mother and baby; “questioners” were more likely to be less confident in safety. It is important to note that respondents were asked about approved maternal vaccines broadly and data were collected before RSV vaccine licensure, WHO recommendation, or availability. As Kenya and other LMICs prepare for potential maternal RSV introduction, it will be important to keep in mind that these types of information can sway perspectives on vaccine safety and demonstrate how decision-makers examined these data to shape their decision.

Social marketing strategies could provide a framework to tailor approaches to the two classes based on the 4 Ps: product, price, placement, and promotion (53,54). “Questioners” were more likely to be seeking care in private facilities, where they likely are paying at least some fee for vaccination and care (compared to lower or no-cost public facilities). Tailored messages for “questioners” should emphasize the seriousness and pervasiveness of RSV in newborns in private facilities in advance of vaccine availability to build knowledge and awareness among women of reproductive age as well as their family and community members. Messages delivered through promotional materials and providers should also communicate the simplicity of RSV vaccination—one dose in the third trimester—and how effective and safe vaccines are at helping mothers protect their newborns. Social marketing has been used effectively to promote childhood and adolescent vaccination in a range of settings and may offer promise for reaching “questioners” where they are and with information that resonates (55–58).

It is unlikely that mAbs will be available in LMIC settings in the near term, at least in public health facilities. Recognizing that “questioners” in our study who express reluctance and distrust in maternal vaccines and RSV disease tend to be private sector facility users, it is possible that private sector availability (at cost) of mAbs is an intermediate strategy to protect children.

Providers in private facilities where mAbs may be available should be equipped to explain the benefits and tradeoffs of maternal vaccination and mAbs to their patients. A recent discrete choice experiment and latent class analysis (43) explored preferences between maternal vaccination and mAbs in the USA where both are available, and found that most respondents (classified as prioritizing effectiveness [51.4%] or duration of protection in RSV season [39.2%]) preferred a maternal vaccine over mAbs, with only those indicating they prioritized no preventive intervention (9.4%) preferring mAbs.

## Limitations and strengths

We acknowledge several important limitations in this analysis. We constrained our LCA model parameters to 12 variables relevant for vaccine hesitancy and acceptance frameworks with unique interpretations; we explored alternative models with more parameters but sought to improve identifiability. Some parameters include related concepts (i.e. safety for mother, safety for baby; injunctive and descriptive norms) and may benefit from further exclusion or consolidation in a subsequent analysis. Boundary issues were noted and examined for safety (mother) and benefits/effectiveness (mother) variables in all models tested, although we retained these variables in the model as it was not inappropriate or unexpected to see consistent responses for these items relating to perceived safety and effectiveness of a maternal vaccine for the mother. We prioritized parsimony when choosing between the two models (2-class, 3-class) given the sufficient but relatively small sample size. We conducted this survey in a facility setting over a relatively short period of time, introducing selection bias into our analysis as we are sampling only individuals seeking care in the facility and not those who are utilizing community-based care or other mechanisms. Social desirability bias is likely.

Despite these limitations, this study has several strengths. This appears to be the first study of its kind to apply latent class analysis for maternal immunization in an LMIC setting. Our exploratory variables and class parameters were grounded in established frameworks and literature, focusing on the most important factors influencing vaccine acceptance and demand. We engaged a diverse range of respondents, including pregnant and postpartum women seeking care in almost all health facility levels across each county, reflecting an array of communities and maternal and newborn care experiences in LMIC settings.

## Conclusion

While most pregnant and breastfeeding women in Nakuru and Mombasa counties express high levels of trust in maternal vaccination for RSV and other diseases, about one in four are reluctant and skeptical about new vaccines in pregnancy. Health providers remain a trusted and important source of information, emphasizing the need to equip providers—especially in private facilities—to navigate new questions about RSV and vaccination, particularly in light of lower RSV awareness and knowledge and safety signals from some trials. Strengthening social awareness of RSV disease and support for maternal vaccination may also help encourage voluntary acceptance of upcoming vaccines. Gaining a better understanding of pregnant and lactating women’s perspectives on community and peer expectations around maternal vaccination is an important first step in shaping health promotion and education strategies for a potential RSVpreF introduction.

Kenya’s national immunization decision-makers are already examining available evidence and considering potential maternal vaccine introduction strategies, setting the stage to be early adopters of this crucial intervention among LMICs. In preparation for such an introduction into the national immunization program, it is essential to understand if and how these vaccines may be viewed by pregnant women and what questions remain to build demand. Our analysis provides one strategy to inform targeted and tailored behavior change activities for each of two cadres—women who are confident about the need, safety, and benefits of maternal vaccines and likely to accept a new maternal RSV vaccine, and women who express ambivalence or even distrust about maternal vaccination.

## Data Availability

Relevant quantitative data are within the paper and its supporting information underlying survey data are available in the primary descriptive paper (https://academic.oup.com/jpids/article/12/12/638/7379696?login=true). Interested parties can request de-identified survey data by contacting the Johns Hopkins University Bloomberg School of Public Health Institutional Review Board requests for access to the underlying study data will be reviewed and approved upon reasonable request.

## Acknowledgements

The authors are sincerely grateful to the pregnant and lactating women who willingly provided their time, experience, and insights for this study, and to the administrators and health officials who facilitated this study’s conduct. The authors also acknowledge the contributions of the Jhpiego Kenya study management team and data collectors who expertly administered surveys and oversaw data collection activities across the two study counties.

## Supporting information

S1 Table. Survey items and response options for variables included in the LCA model a Responses were dichotomized to strongly agree/agree and disagree/strongly disagree/don’t know for analysis; b RSV = Respiratory syncytial virus

